# Poverty associated with the environmental contamination of gastrointestinal parasites in the Southern United States

**DOI:** 10.1101/2023.01.10.23284404

**Authors:** Christine Crudo Blackburn, Sally Mingshuang Yan, David McCormick, Lauren Nicholas Herrera, Roumen Borilov Iordanov, Mark Daniel Bailey, Maria Elena Bottazzi, Peter J. Hotez, Rojelio Mejia

## Abstract

**Background:** Gastrointestinal parasites are generally associated with lower-income countries in tropical and subtropical areas, but they are also prevalent in low-income and extreme low-income communities in the Southern United States. To date, studies characterizing the epidemiology of gastrointestinal parasites in the United States are limited, resulting in little comprehensive understanding of the challenge. This study investigates the environmental contamination of gastrointestinal parasites in the Southern United States by determining the contamination rate and burden of each parasite in five low-income communities.

**Methods:** A total of 499 soil samples of approximately 50g were collected from public parks and private residences in Alabama, Louisiana, Mississippi, South Carolina, and Texas. A novel technique utilizing parasite floatation, filtration, and bead-beating was applied to concentrate and extract parasite DNA from samples and detected via multi-parallel qPCR.

**Findings:** qPCR detected *Blastocystis* spp (19.0%), *Toxocara cati* (6.01%), *Toxocara canis* (3.61%), *Strongyloides stercoralis* (2.00%), *Trichuris trichiura* (1.80%), *Ancylostoma duodenale* (1.42%), *Giardia intestinalis* (1.40%), *Cryptosporidium* spp (1.00%), *Entamoeba histolytica* (0.201%), *and Necator americanus* (0.200%). Overall parasite contamination rates varied significantly between communities: Western Mississippi (46.88%); Southwestern Alabama (39.62%); Northeastern Louisiana (28.24%); Southwestern South Carolina (27.03%); and South Texas (6.93%) (p < 0.0001). *Toxocara cati* DNA burdens were greater in communities with higher poverty rates, including Northeastern Louisiana (50.57%) and Western Mississippi (49.60%) compared to Southwestern Alabama (30.05%) (p = 0.0011).

**Interpretation:** This study demonstrates the environmental contamination of parasites and their relationship with high poverty rates in communities in the Southern United States.

**Funding:** This work was supported by the Maternal and Infant Environmental Health Riskscape (MIEHR) Center of Excellence on Environmental Health Disparities Research, NIMHD grant #P50 MD015496

**Research in context:** *Evidence before this study:* Several research articles on parasites in the Southern USA were used to determine the extent of parasitosis in the selected regions (Mckenna et al, *Am J Trop Med Hyg* 2018; Singer et al, *Am J Trop Med Hyg* 2020; Bradbury et al, *Emerg Infect Dis* 2021). Criteria used included terms hookworm, soil-transmitted helminths, protozoa, and parasites in the United States. The most recent parasite prevalence studies indicate areas with 62.9% *Blastocystis*, 34.5% hookworm (*Necator americanus*), 16.5% *Strongyloides stercoralis*, 5.2% *Toxocara*, 2.9% *Cryptosporidium*, 2.3% *Giardia intestinalis*, and 1.8% *Entamoeba histolytica* from human serum or stool samples. These studies focus on human parasite infections, but there are no current published studies on environmental parasite surveys in the Southern US. The parasites in this study all have part of their life-cycle in soil and can directly infect humans living in these areas.

*Added value of this study:* Our work expands the current understanding of prevalence of parasites in the Southern US soil from built environments. We further correlate the type of parasites and their intensity of soil contamination with higher poverty rates.

*Implications of all the available evidence:* These parasitic infections represent a major source of contamination in the built environments and their association with poverty. This study will help focus public policy on the potential risks that environmental parasites have on human health. It further describes the high contamination rates in the USA, a high-income country.

## Introduction

Neglected infections of poverty are infectious diseases that disproportionately affect marginalized communities[1]. Many gastrointestinal parasites are among these neglected infections of poverty, specifically *Ascaris lumbricoides, Cryptosporidium* spp. *Entamoeba histolytica, Giardia intestinalis, Strongyloides stercoralis*[1]. The symptoms caused by these gastrointestinal parasites vary but generally include diarrhea, anemia, and malnutrition [2]. As a result, they can cause delays in cognitive and physical development in childhood, reinforcing the cycle of poverty [3, 4].

Despite their clinical and societal significance, there are limited studies to characterize the epidemiology of gastrointestinal parasites in the United States. Most systematic, high-quality studies of their prevalence were conducted from 1942 to 1982, meaning that current information is limited [5]. Furthermore, the latter studies concluded that endemic transmission of soil-transmitted helminths persisted. More recently, the 1999-2004 and 2009-2010 National Health and Nutrition Examination Survey (NHANES) analyzed the prevalence of some gastrointestinal parasites, including *Toxocara canis* and *Toxocara cati* [6]. Other recent studies characterized the population prevalence of several parasites in rural Alabama, peri-urban Texas, and among Latin American immigrants in Washington, D.C. [2, 7, 8].

To address the limited existing data, environmental sampling provides an opportunity for broader epidemiological studies. This approach may indicate population-level prevalence and potential for transmission and, thus, demonstrate a need for further study and funding. Most environmental studies of parasites are wastewater-based epidemiology studies of protozoa such as *Cryptosporidium* spp. and *Giardia intestinalis* [9, 10]. Such studies largely exclude helminths, which are primarily transmitted through the soil and whose life cycles entail defecation into the soil by humans or animals followed by ingestion from the soil by humans [2]. Additionally, protozoa and heterokonts are also present in the soil and, thus, could possibility be transmitted through it [11]. Some studies have aimed to detect *Toxocara* spp. and other soil-transmitted helminths in soil samples [12, 13]; however, the two such studies conducted in the United States sampled sewage sludge rather than soil [12, 14]. Furthermore, these studies utilized conventional microscopy-based detection of helminth eggs, which are subjective and inaccurate. In contrast, qPCR-based molecular detection methods are more sensitive, less labor intensive, and less time consuming [15].

This project utilizes the molecular detection of 11 gastrointestinal parasites in soil samples in five extreme low-income communities in the Southern United States. The five communities are in the highest quintile for poverty rate in the United States and include cities and counties in Southwestern South Carolina, Northeastern Louisiana, South Texas, Western Mississippi, and Southwestern Alabama. It characterizes the contamination rate and burden of each parasite in each community as possible indicators of the prevalence among human populations and potential for endemic transmission in the United States. Furthermore, it examines the association between parasite contamination rates and poverty in the United States (Figure 1).

**Figure 1.**
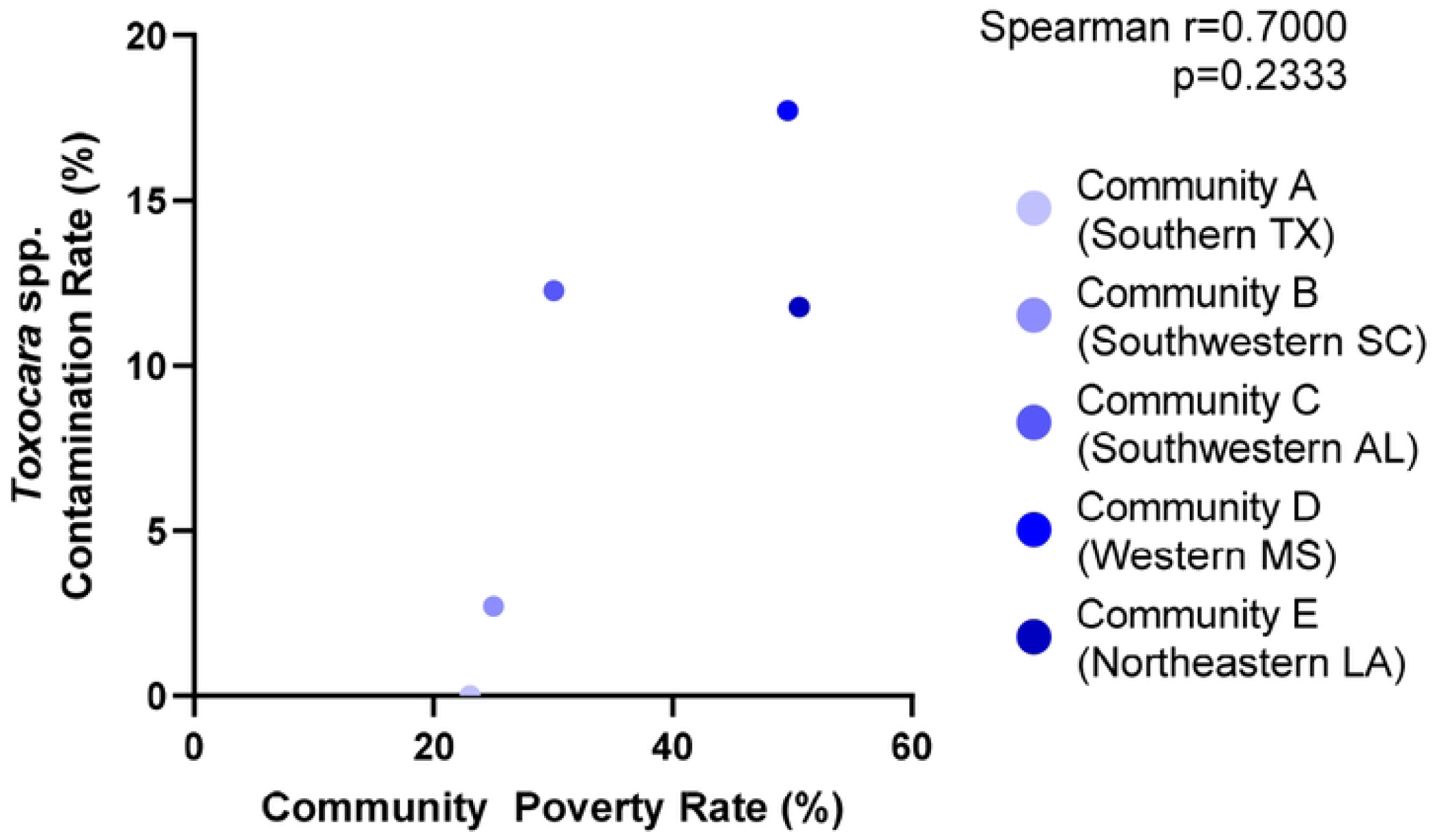
Map of communities and their respective poverty rates. Quintiles of poverty rates were retrieved from the 2018 American Community Survey and displayed.

This paper reports the results from a research study of 499 soil samples, for which DNA extraction and qPCR testing have been carried out. Among this subset of samples, several parasites—including *Blastocystis* spp (19.0%), *Toxocara cati* (6.01%), *Toxocara canis* (3.61%), *Strongyloides stercoralis* (2.00%), *Trichuris trichiura* (1.80%), *Ancylostoma duodenale* (1.42%), *Giardia intestinalis* (1.40%), *Cryptosporidium* spp (1.00%), *Entamoeba histolytica* (0.201%), *and Necator americanus* (0.200%)—were detected. Additionally, parasite contamination rates were significantly associated and correlated with poverty rates.

## Methods

### Study Design and Sample Collection

Five cities or counties in five states in the Southern United States were selected as study communities. These included: a county located in Southwestern South Carolina; a city in Northeastern Louisiana; a city in South Texas; a city in Western Mississippi; and a county in Southwestern Alabama. Criteria for the selection of communities included high poverty rate, low household median income, and rural status (Table 1). Only the community located in South Texas is classified as urban and the decision to include this community was based on its proximity to large agricultural production and the U.S.-Mexico border. A total of approximately 100 samples, over four to six sites, were collected from each community. Collection sites in each community were a combination of public parks and private residences.

**TABLE 1.** Communities selected for the study with their corresponding poverty rates (%), annual household median incomes ($), and populations (# of people).*

| Community | Location | Median Household Income (\$) | Poverty Status (%) | Population (#) |
| --- | --- | --- | --- | --- |
| A | South TX | \$47,279.00 | 23.05% | 141,968 |
| B | Southwestern SC | \$24,560.00 | 25.01% | 9,024 |
| C | Southwestern AL | \$27,237.00 | 30.05% | 10,681 |
| D | Western MS | \$20,857.00 | 49.60% | 2,004 |
| E | Northeastern LA | \$17,801.00 | 50.57% | 2,753 |

\* Poverty rates, household median incomes, and populations were retrieved from the 2019 American Community Survey conducted by the U.S. Census Bureau.

For each sample, approximately 50 grams of soil was collected by scraping a 50 mL conical centrifuge tube along the surface of the soil in different locations within the site. These samples were transported and stored at 4°C.

### Parasite Floatation and Filtration

A novel DNA concentration technique using parasite flotation and filtration was used to concentrate parasite DNA from soil samples before DNA extraction. The mass of the samples— ranging from approximately 5 to 80 grams—were determined and recorded prior to extraction. If more than 80 grams of soil were collected for a sample, 50 grams was used for parasite floatation and filtration followed by DNA extraction, and the remainder was reserved.

Each sample was divided in half between two 50 mL conical centrifuge tubes. To wash macro-scale debris from the soil samples, PBS (Alfa Asesar, Ward Hill, MA) with 0.05% TWEEN (Sigma-Aldrich, St. Louis, MO) was added to the 50 mL for each sample. The samples were vortexed for 5 minutes, centrifuged at 500 g for 5 minutes, and the supernatant containing the debris was discarded.

To float helminth eggs and larva, as well as protozoa, 10 mL of a 35.6% NaNO_3_ solution (Vedco, St. Joseph, MO) with a specific gravity of 1.25-1.30 was added to the pellet in each conical centrifuge tube. The solution was vortexed for 5 minutes and centrifuged for 5 minutes at 500 g.

The supernatant for each sample containing the floated parasites was then transferred to a filtration apparatus. The filtration apparatus consisted of a 50 mL syringe attached to a 50 mm syringe filter containing a nitrocellulose filter with 3 μm pores (MilliporeSigma, Burlington, MA), which is small enough to retain all parasites subsequently tested for. The filtration apparatus was attached to a vacuum manifold, which was, in turn, attached to a two stage rotary vane vacuum pump (ELITech, Puteaux, FR). Filtration with a vacuum pressure of as low as 25 microns Hg was performed for a duration of 60 minutes.

### DNA extraction

The MP Fast SpinKit for Soil (MP Biomedicals, Santa Ana, CA) was utilized with a modified protocol to extract DNA from parasites retained on the nitrocellulose filter. The modifications entailed preliminary steps to lyse parasite eggs. In brief, filters were transferred with tongue blades to a lysing solution. The solution contained a lysing matrix with ceramic, glass, and silica beads; 978 μL sodium phosphate buffer; 122 μL MT buffer; and an internal control DNA sequence subsequently used to confirm successful extraction. Heat disruption at 90°C for 10 minutes in a dry bath incubator followed by mechanical disruption by bead beating in the MP FastPrep 34-5G disruptor (MP Biomedicals, Santa Ana, CA) on speed 6 for 40 seconds was used to break open parasite eggs and lyse cells. Subsequent steps followed the standard protocol of the MP Fast SpinKit for Soil to extract DNA.

### qPCR Testing for Parasite DNA

The DNA extracted from samples were tested for *Ascaris lumbricoides, Ancylostoma duodenale, Toxocara cati, Toxocara canis, Cryptosporidium* species, *Entamoeba histolytica, Giardia intestinalis, Necator americanus, Strongyloides stercoralis, Trichuris trichiura*, and *Blastocystis* subtypes using quantitative PCR.

To test for each parasite, a 7 μL reaction mixture was prepared for each sample. The reaction mixture consisted of 5 μL TaqMan® Fast Advanced Master Mix (Applied Biosystems, Foster City, CA) with previously published forward primers (900 nM final concentration) (Applied Biosystems, Foster City, CA), reverse primers (900 nM final concentration) (Applied Biosystems, Foster City, CA), and FAM probe with a minor groove binder and non-fluorescent quencher (100 nM final concentration) (Applied Biosystems, Foster City, CA) for each parasite (Table 2). Additionally, 2 μL of extracted DNA was added to each reaction mixture.

**TABLE 2.**
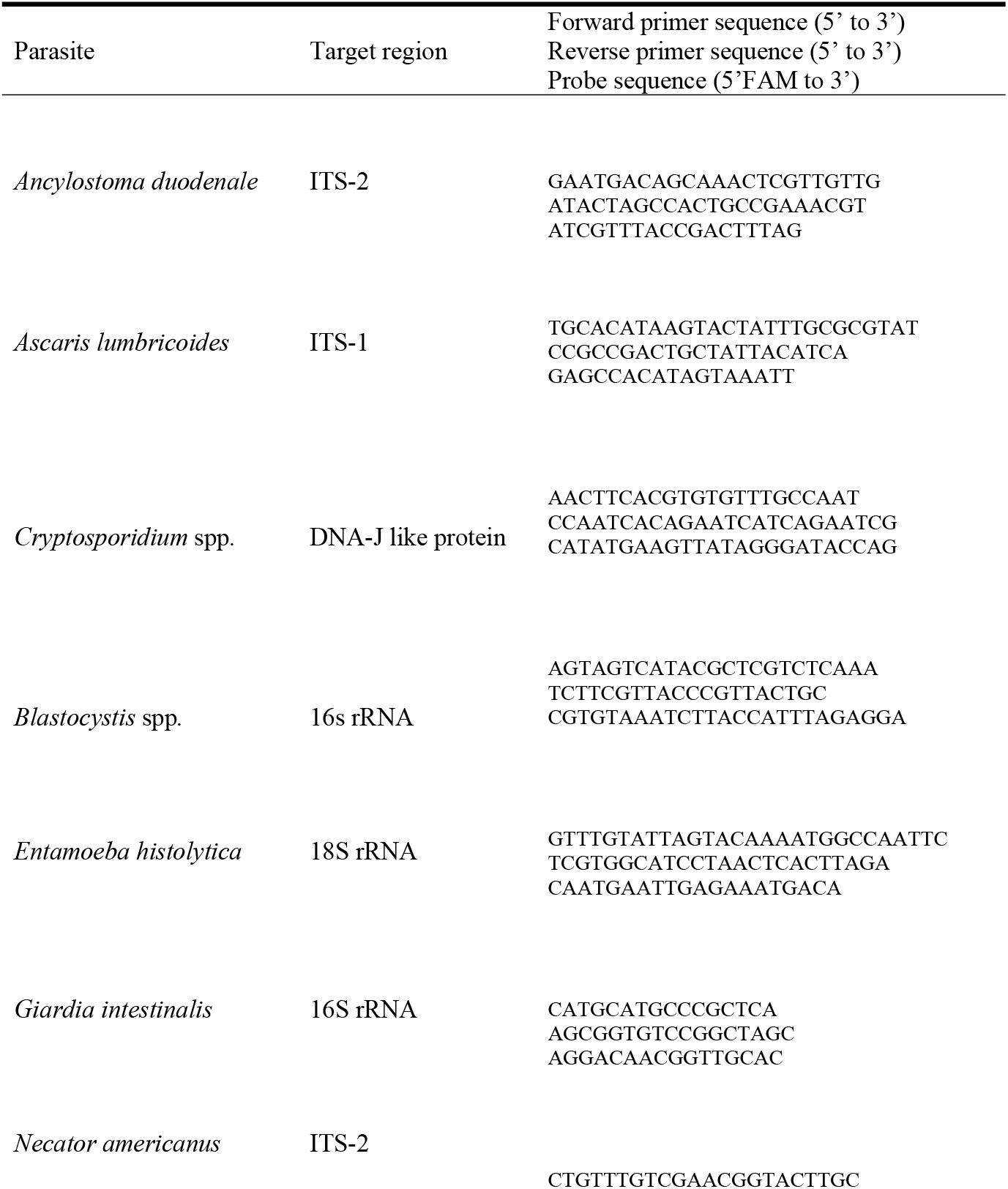
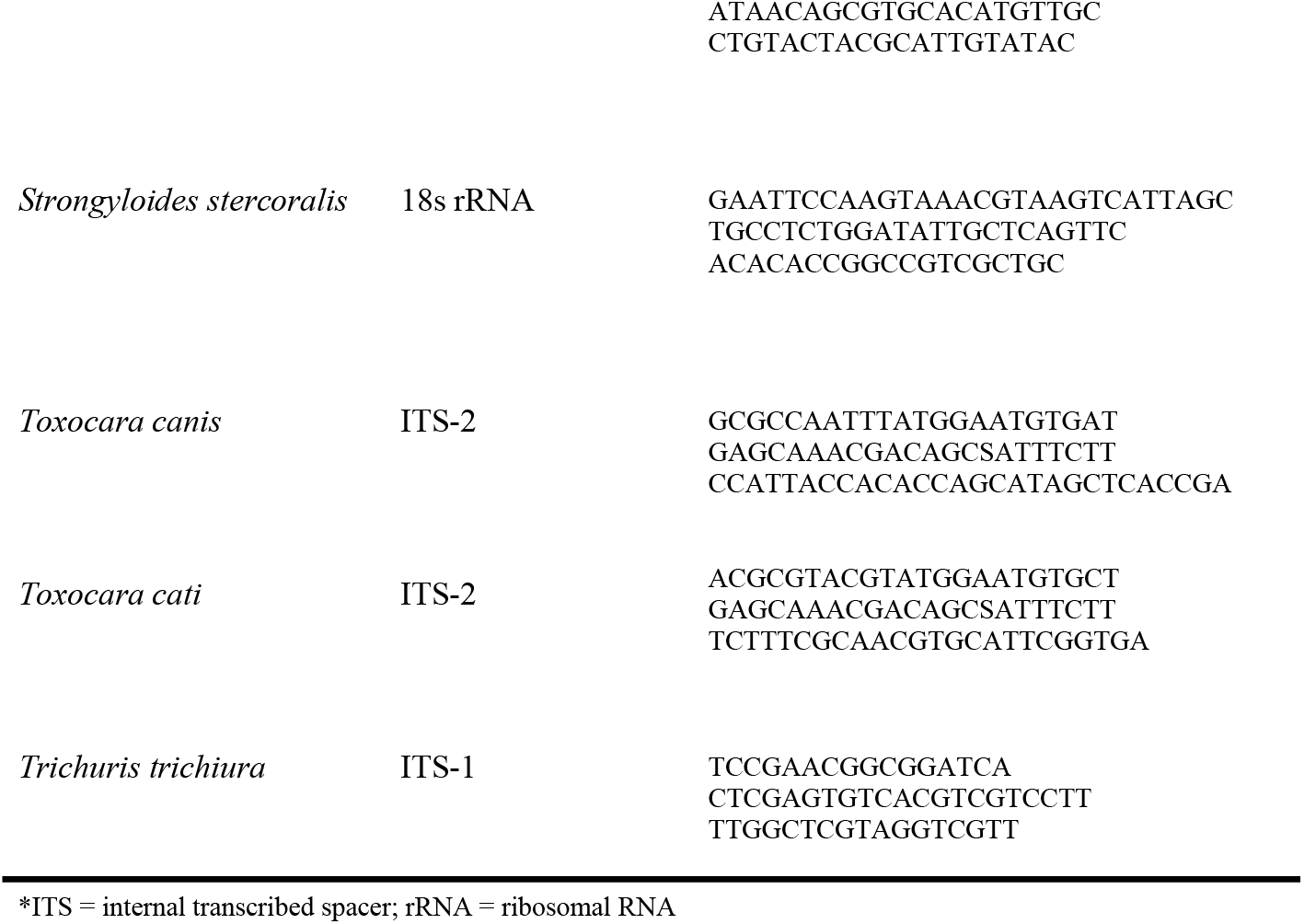
Target regions, primer sequences, and probe sequences by parasites for DNA amplification.*

A parasite-plasmid standard curve of 10-fold dilutions was generated for each parasite to serve as a positive control and to allow the quantification of the concentration of parasite DNA. Nuclease free water was used as a negative control.

The Fast Chemistry protocol for a 7 μL reaction volume was performed on the ABI Vii-A7, QuantStudio™ 3, or QuantStudio™ 7 Real-Time PCR systems with a hold stage and 40 cycles of amplification (Applied Biosystems, Foster City, CA) (Table 3). Results were analyzed on the QuantStudio™ Design and Analysis v2.6.0 software. Based on a previously established dynamic range using parasite-plasmid standards, samples were considered positive for Ct < 40.[16]

**TABLE 3.**
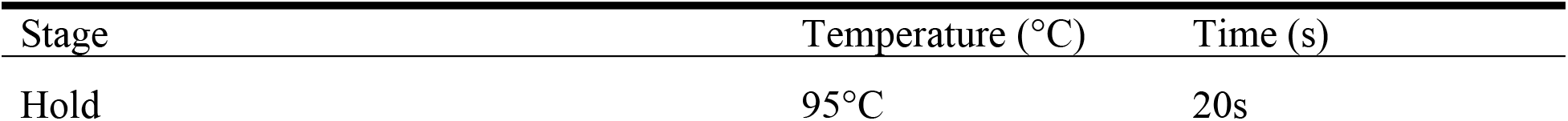
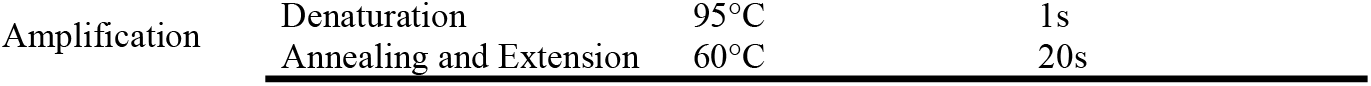
Run method for Fast Chemistry protocol on Real-Time PCR systems.

| Stage | Temperature (°C) | Time (s) |
| --- | --- | --- |
| Hold | 95°C | 20s |

|  |  |  |  |
| --- | --- | --- | --- |
| Amplification | Denaturation | 95°C | 1s |
|  | Annealing and Extension | 60°C | 20s |

#### Data Analysis

The contamination rate and median parasite burdens were noted for each parasite in each community, and socioeconomic indicators were recorded for each community. The contamination rate was calculated as the percentage of positive samples per the total number samples tested. The parasite burden was defined as the concentration of the target DNA sequence for each parasite in fg/μL quantified using the standard curve and normalized by soil sample mass. Socioeconomic indicators for each community including poverty rate, median household income, and GDP per capita were obtained from the 2018 American Community Survey conducted by the U.S. Census Bureau.

Using these variables, statistical analysis was performed on GraphPad Prism v9.4.0(GraphPad Software, La Jolla, CA). Specifically, Chi-squared tests were conducted to examine the association of socioeconomic indicators of communities with their parasite contamination rates. Kruskal-Wallis tests were utilized to examine the association of socioeconomic indicators of communities with their median parasite burdens. Spearman’s rank correlation tests were applied to investigate the correlation between socioeconomic indicators of communities with parasite contamination rates or median parasite burdens.

## Results

### Environmental Contamination of Parasites

Four hundred ninety-nine (499) samples were tested for *Ancylostoma duodenale, Cryptosporidium* species, *Entamoeba histolytica, Giardia intestinalis, Necator americanus, Strongyloides stercoralis, Toxocara canis, Toxocara cati, Trichuris trichiura*, and *Blastocystis* spp through qPCR. The findings are summarized in Table 4. For the cohort overall, *Blastocystis* spp. was the parasite with the highest environmental contamination rate (19.0%, 95/499). Among the communities, Community D in Western MS had the highest contamination rate for *Blastocystis* spp. (28.1%, 27/96), followed by Community C in Southwestern AL (23.6%, 25/106), Community B in Southwestern SC (22.5%, 25/111), Community E in Northeastern LA (14.1%, 12/85), and Community D South TX (5.94% 6/101).

**TABLE 4.** Contamination rates by samples and by sites for each parasite detected overall and by community.*

| Parasite | Contamination Rate by Samples | Contamination Rates by Sites |
| --- | --- | --- |
| *Blastocystis* spp. |  |  |
| Overall | 19.0% (95/499) | 73.0% (27/37) |
| Southwestern SC | 22.5% (25/111) | 100% (5/5) |
| Northeastern LA | 14.1% (12/85) | 55.6% (5/9) |
| South TX | 5.94% (6/101) | 57.1% (4/7) |
| Western MS | 28.1% (27/96) | 72.7% (8/11) |
| Southwestern AL | 23.6% (25/106) | 100% (5/5) |
| *Toxocara cati* |  |  |
| Overall | 6.01 (30/499) | 37.8% (14/37) |
| Southwestern SC | 0.90% (1/111) | 20.0% (1/5) |
| Northeastern LA | 9.41% (8/85) | 44.4% (4/9) |
| Western MS | 10.4% (10/96) | 45.5% (5/11) |
| Southwestern AL | 6.01% (11/95) | 80.00% (4/5) |
| *Toxocara canis* |  |  |
| Overall | 3.61% (18/499) | 24.3% (9/37) |
| Southwestern SC | 2.70% (2/111) | 40.0% (2/5) |
| Northeastern LA | 2.35% (2/85) | 22.2% (2/9) |
| Western MS | 9.38% (9/96) | 36.4% (4/11) |
| Southwestern AL | 3.77% (4/106) | 20.0% (1/5) |
| *Strongyloides stercoralis* |  |  |
| Overall | 2.00% (10/499) | 13.5% (5/37) |
| Southwestern SC | 5.41% (6/111) | 40.0% (2/5) |
| Western MS | 3.13% (3/96) | 18.2% (2/11) |
| Southwestern AL | 0.943% (1/106) | 20.0% (1/5) |
| *Trichuris trichiura* |  |  |
| Overall | 1.80% (9/499) | 13.5% (5/37) |
| Northeastern LA | 1.18% (1/85) | 11.1% (1/9) |
| Western MS | 6.25% (6/96) | 27.3% (3/11) |
| Southwestern AL | 1.89% (2/106) | 20.0% (1/5) |
| *Ancylostoma duodenale* |  |  |
| Overall | 1.42% (7/494) | 13.5% (5/37) |
| SouthwesternSC | 0.909% (1/110) | 20.00% (1/5) |
| Northeastern LA | 2.381% (2/84) | 22.2% (2/9) |
| South TX | 0.990% (1/101) | 14.3% (1.7) |
| Western, MS | 3.13% (3/96) | 9.09% (1/11) |
| *Giardia intestinalis* |  |  |
| Overall | 1.40% (7/499) | 10.8% (4/37) |
| Southwestern SC | 1.80% (2/111) | 40.0% (2/5) |
| Western MS | 4.17% (4/92) | 9.09% (1/11) |
| Southwestern AL | 0.943% (1/106) | 20.0% (1/5) |
| *Cryptosporidium* spp. |  |  |
| Overall | 1.00% (5/493) | 5.40% (2/37) |
| South TX | 0.990% (1/101) | 0.00% (0/7) |
| Southwestern AL | 3.77% (4/102) | 40.0% (2/5) |
| *Entamoeba histolytica* |  |  |
| Overall | 0.201% (1/497) | 0.00% (0/37) |
| Southwestern AL | 0.952% (1/105) | 0.00% (0/5) |
| *Necator americanus* |  |  |
| Overall | 0.200% (1/499) | 0.000% (0/37) |
| Western MS | 1.04% (1/96) | 0.00% (0/11) |

\*Contamination rate by sample is calculated as the percentage of positive samples per all samples tested for a parasite. Positive samples/all samples tested displayed in parentheses. Contamination rate by site is calculated as the percentage of positive sites per all sites. Positive sites/all sites displayed in parentheses. If a site has one positive sample, it is considered positive for the parasite.

The zoonotic soil-transmitted helminths *Toxocara cati* (8.62%, 43/499) and *Toxocara canis* (6.01%, 30/499) had the next highest contamination rates. For *Toxocara cati*, the contamination rates for the different communities in which *Toxocara cati* was detected respectively were: Community D in Western Mississippi (10.4%, 10/96), Community C in Southwestern Alabama (10.4%, 11/106), Community E in Northeastern Louisiana (9.41%, 8/85), and Southwestern South Carolina (0.90% (1/111). Soil samples positive for *Toxocara canis* were found in: Community D in Western Mississippi (9.38%, 9/96), Community C in Southwestern Alabama (3.77%, 4/106), Community B in Southwestern South Carolina (2.70%, 2/111), and Community A Northeastern Louisiana (2.35%, 2/85).

Other soil-transmitted helminths detected in the environment included *Strongyloides stercoralis* (2.00%, 10/499), T*richuris trichiura* (1.80%, 9/499), *Ancylostoma duodenale* (1.42%, 7/499), and Necator americanus (0.200%, 1/499) Six of the 10 positive samples for *Trichuris trichiura* were collected from the community in Western Mississippi (6.25%, 6/96). *Necator americanus* was only detected in the community in Western Mississippi (1.04%, 1/96) while *Strongyloides stercoralis* was found most commonly in the community in Southwestern South Carolina (5.41%, 6/111).

The protozoa *Giardia intestinalis (*1.40%, 7/499), Entamoeba histolytica (0.201%, 1/497), and *Cryptosporidium* spp. (1.00%, 5/493) were also detected. For *Giardia intestinalis*, the highest environmental contamination rate was found in the community in Western Mississippi (4.17%, 4/96) whereas two positive samples each were found in Southwestern South Carolina (1.80%, 2/111) and one positive sample was found in Southwestern Alabama (0.943%, 1/106). However, for *Cryptosporidium* spp., Southwestern Alabama had the highest environmental contamination rate (3.77%, 4/102), and one additional positive soil sample was collected from South Texas (0.99%, 1/101). The only sample positive for *Entamoeba histolytica* was collected in Southwestern Alabama (0.952%, 1/105).

### Relation of Environmental Contamination of Parasites to Poverty

To characterize the relation between the environmental contamination of parasites and community poverty rates, the overall parasite contamination rate for each community was identified and utilized as an index despite the biological differences between the different parasites. Positive samples for any parasite tested were considered positive for this index. The relationship between poverty and parasite contamination is shown in Figure 2 (p<0.0001).

**Figure 2.**
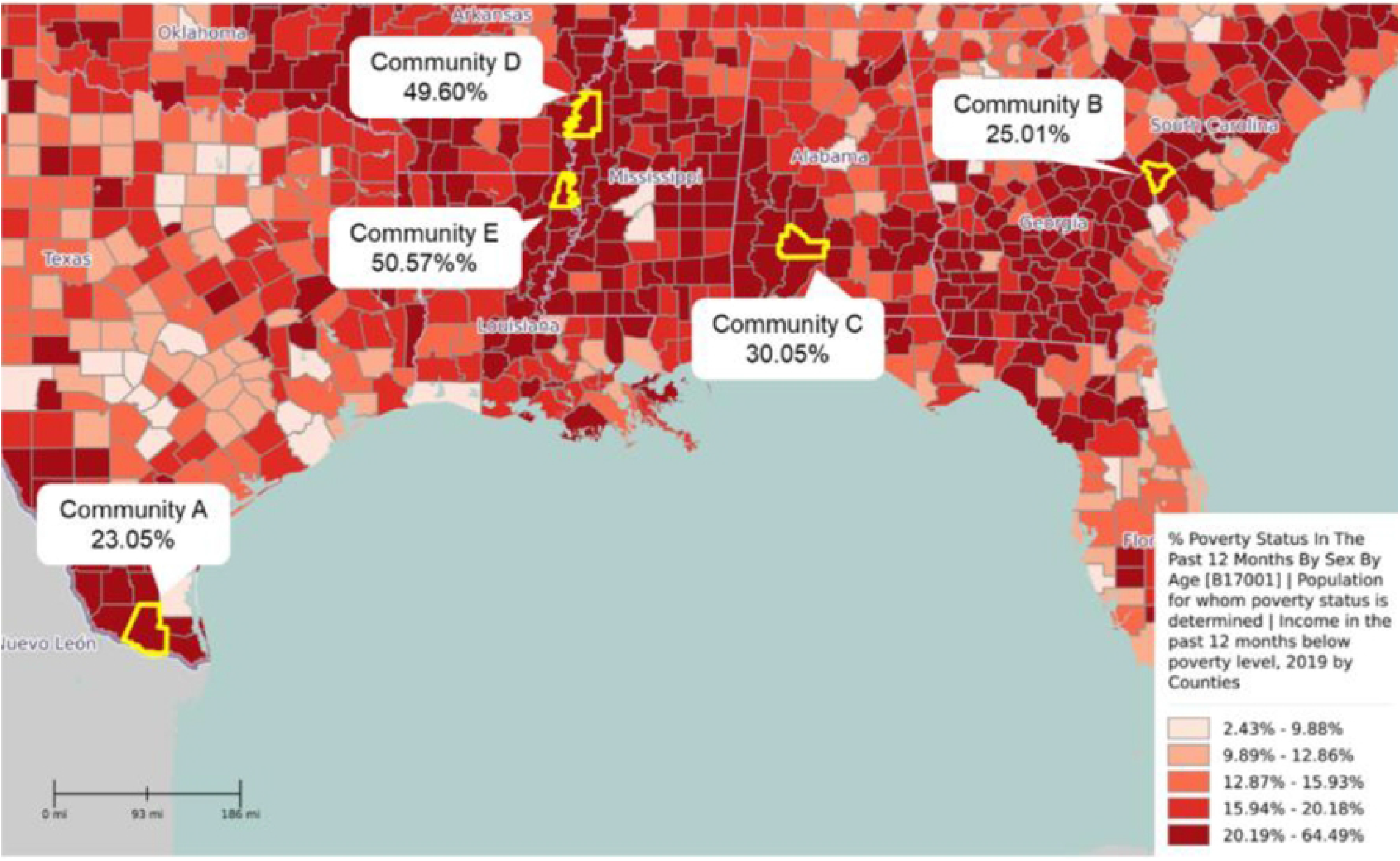
Overall parasite contamination rate (%, black dots) and poverty rates (%, blue bars) by community. Communities with higher poverty rates had higher significantly parasite contamination rates (p<0.0001). Contamination rate was calculated as # of positive samples/total # of samples tested x 100%. Samples positive for any parasite tested were considered positive for overall parasite contamination rate. Community poverty rates were obtained from the 2019 American Community Survey by U.S. Census Bureau.

Furthermore, the community in Western Mississippi, which had one of the highest poverty rates of the communities studied (49.60%), featured the highest overall parasite contamination rate (46.88%). The parasite contamination rate generally decreased as poverty rates decreased across the communities, where the communities with the lowest poverty rate—Southwestern South Carolina (25.01%) and South Texas (23.05%)—also had the lowest overall parasite contamination rates of 27.91% and 7.92% respectively. However, Northeastern Louisiana proved an outlier for this trend with the highest poverty rate (50.57%) but an overall parasite contamination rate of only 28.24%.

Examining specific parasites, the environmental contamination rates for *Toxocara* spp, which includes both *Toxocara cati* and *Toxocara canis*, displayed a similar relation to community poverty rates. The communities in the study had significantly different *Toxocara* spp. contamination rates (p<0.0001), and communities with higher poverty rates had higher contamination rates for *Toxocara* spp (Figure 3). South Texas with the lowest poverty rate of 23.05%, did not have any soil samples positive for either *Toxocara cati* or *Toxocara canis*. Northeastern Louisiana served as an outlier again with the highest poverty rate (50.57%) but the third highest *Toxocara* spp. contamination rate (11.76%). This relation between *Toxocara* spp. contamination rates and community poverty rates was further demonstrated by the positive correlation identified (r_s_=0.7000) (Figure 4).

**Figure 3.**
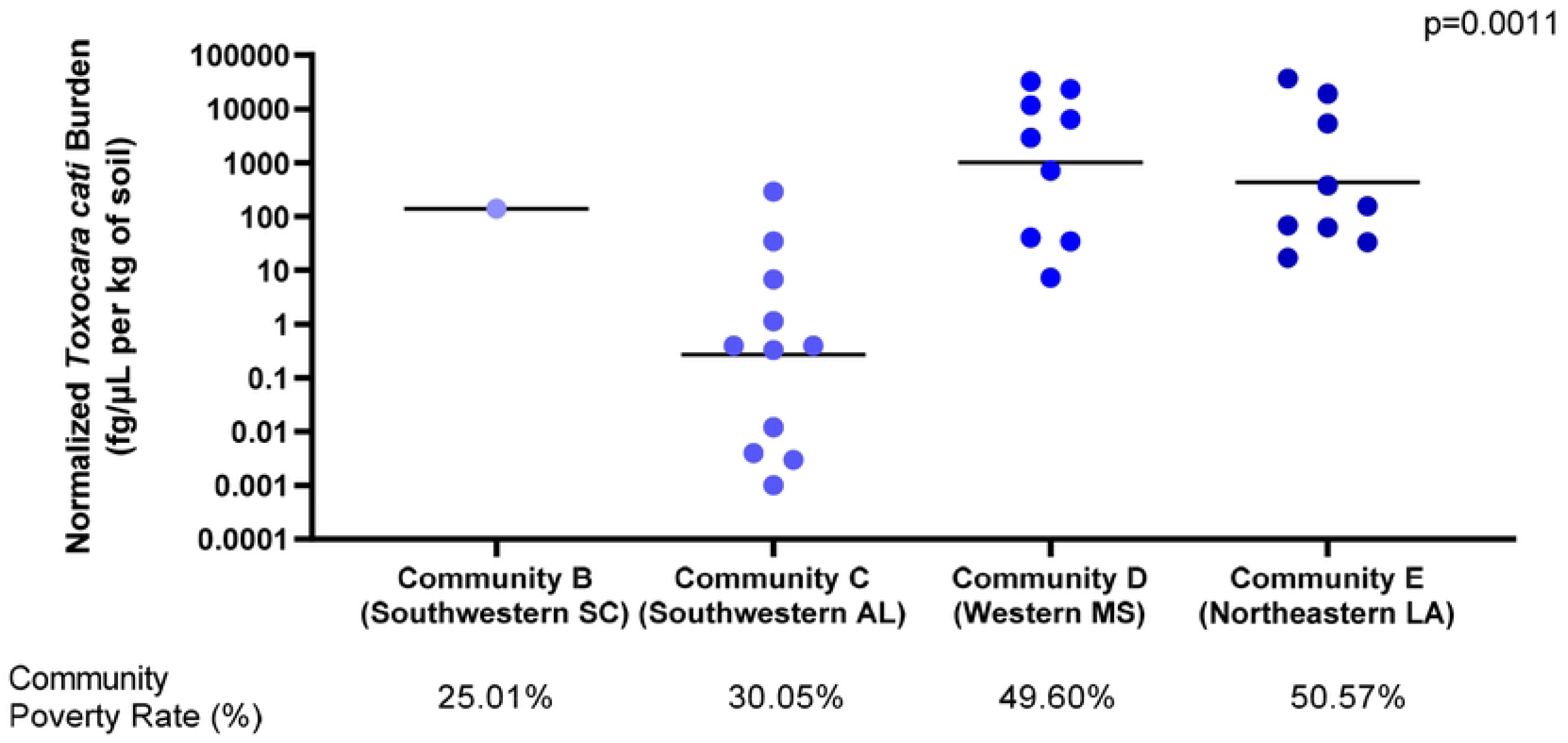
Toxocara spp. contamination rate (%, blue bars) and poverty rates (%, black dots) by community. Communities with higher poverty rates had higher significantly parasite contamination rates (p<0.0001). Contamination rate was calculated as # of positive samples/total # of samples tested x 100%. Samples positive for Toxocara cati or Toxocara canis was considered positive for the Toxocara spp. contamination rate. Community poverty rates were obtained from 2019 American Community Survey by U.S. Census Bureau.

**Figure 4.**
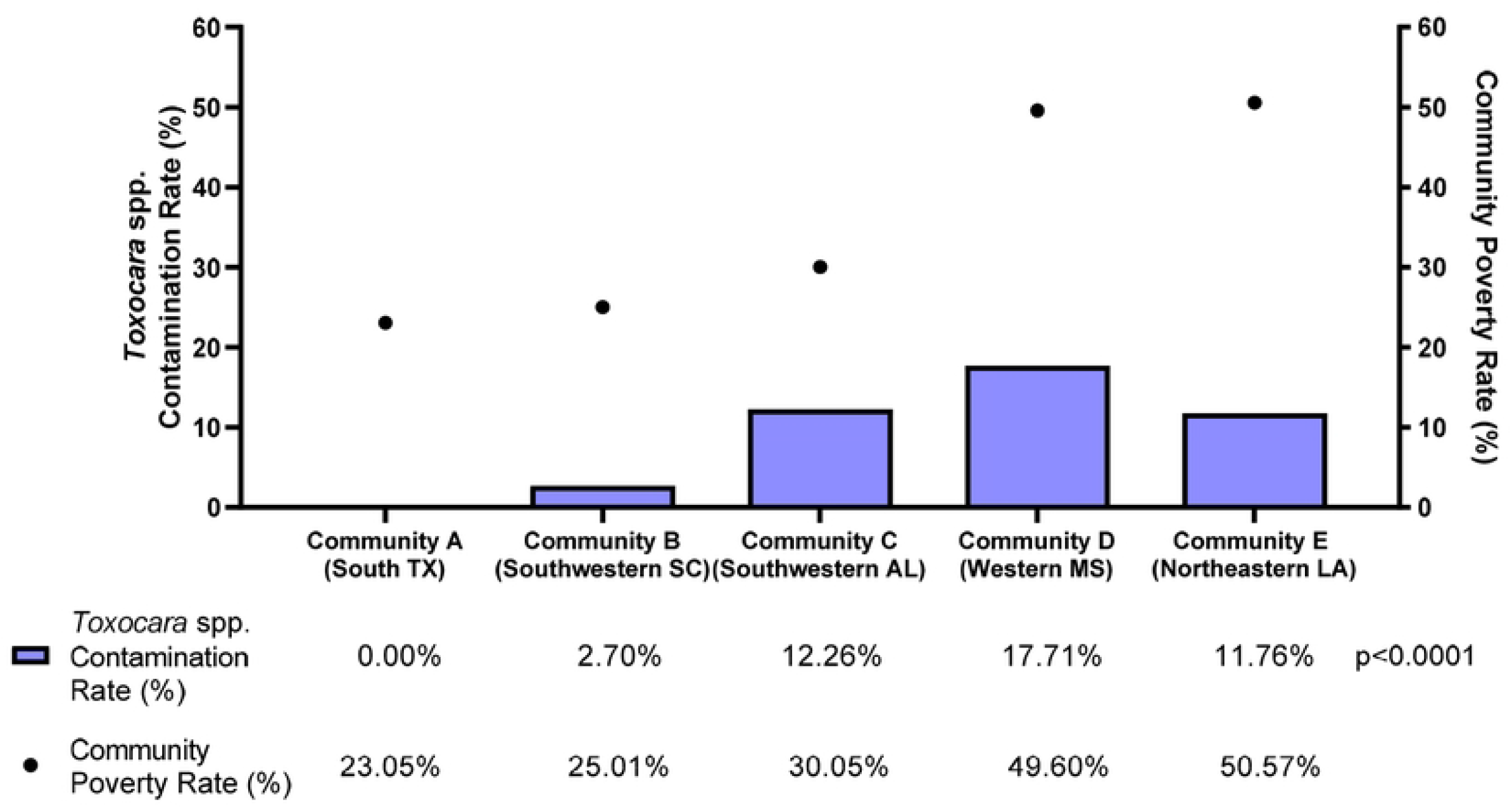
Correlation of *Toxocara* spp. contamination rates (%) with community poverty rates (%). There was a moderately positive correlation between *Toxocara* spp. contamination rates (r_s_=0.7000), but this correlation was not significant (p=0.2333). Contamination rate was calculated as # of positive samples/total # of samples tested x 100%. Samples positive for *Toxocara cati* or *Toxocara canis* are considered positive for *Toxocara* spp. contamination rate. Community poverty rates were obtained from the 2018 American Community Survey by U.S. Census Bureau.

Regarding parasite burdens, quantified as the concentrations of the parasite DNA normalized to the mass of the soil sample, a significant difference was characterized between *Toxocara cati burdens* in the different communities in which it was detected (p=0.0002) (Figure 3). Specifically, the median normalized burdens were 1808 fg/μL of DNA per kg of soil and 155 fg/μl of DNA per kg of soil for Western Mississippi and Northeastern Louisiana, respectively— the two communities with the highest poverty rates of 49.60% and 50.57%, respectively. In contrast, Southwestern Alabama, with a community poverty rate of 30.05%, had a median normalized *Toxocara cati* burden of 0.362 fg/μL per kg of DNA.

To examine one possible confounding factor for the relation between poverty and parasites, the environmental contamination rates and burdens were also compared between public parks and private residences. However, the overall parasite contamination rate (p=.4583) and the contamination rate of *Toxocara* spp. (p=0.8449) were not significantly different between public and private sampling sites. Furthermore, the burden of Toxocara cati did not exhibit any significant difference (p=0.8351) between public parks and private residences.

## Discussion

### Environmental Contamination of Parasites

*Blastocystis* spp., the parasite with the highest environmental contamination rate (19.0%) in this study, is also the most common human parasite in the United States. Large-scale studies of the epidemiology of *Blastocystis* spp. in 2000 and 2004 identified the prevalence as 11-23% among the American population [17, 18]. Furthermore, using more sensitive molecular detection methods, the prevalence was as high as 62.8% in certain rural, low-income communities.^9^ However, there are no other studies of *Blastocystis* spp. in environmental samples in the United States to date. While the pathogenicity of *Blastocystis* spp. in immunocompetent individuals is debated–-some studies demonstrate its association with non-specific gastrointestinal symptoms and others repudiate its association–-the environmental contamination rate of *Blastocystis* spp. could serve as an indicator of overall fecal-oral contamination in the environment [18, 19].

In contrast to *Blastocystis* spp., many studies have characterized the environmental presence of the other unicellular eukaryotic parasites detected in the study—*Giardia intestinalis* (1.40%) and *Cryptosporidium* spp. (1.00%). Studies of the presence of *Giardia intestinalis* and *Cryptosporidium* spp. in water have been conducted both in low-income and high-income settings.[9, 10] Several such studies have characterized the contamination of *Cryptosporidium* spp. or *Giardia intestinalis* in wastewater or surface water in the United States [20, 21].

However, none of these studies examined soil contamination. Although both *Giardia intestinalis* and *Cryptosporidium* spp. are primarily waterborne protozoa, they are also present in the soil [11]. Furthermore, Dai and Boll (2003) demonstrated that their ova attach to soil particles even in aquatic environments [22]. Thus, soil may serve as an additional route of transmission for *Giardia intestinalis* and *Cryptosporidium* spp.

Of the soil-transmitted helminths, *Toxocara cati* (8.62%) and *Toxocara canis* (6.01%) are the most prevalent pathogenic parasites found in this study. Toxocariasis—as both visceral larva migrans and ocular larva migrans—caused by either of these parasites is 1 of 6 Neglected Parasitic Infections in United States designated by CDC.[23] Among human populations, recent NHANES surveys have shown varying prevalence from 5.1% to 13.9%.[24] *Toxocara* spp. are also some of the few parasites with previous characterization in environmental studies [12, 13]. Since domestic cats and dogs are the primary hosts of these parasites, their detection in the soil in the communities may not be indicative of the prevalence in human populations [23]. Nonetheless, as zoonotic pathogens, environmental studies are particularly important since their life cycle requires maturation in the soil prior to transmission to humans [25].

The soil-transmitted helminths *Strongyloides stercoralis* (3.610.246%), *Trichuris trichiura* (2.00%), *Ancylostoma duodenale* (1.42%), and *Necator americanus* (0.200%), were also detected. These four parasites are some of the most common soil-transmitted helminths globally [26]. In the United States, a previous large-scale study in Kentucky in 1982 found prevalences of 12.6% for *Trichuris trichiura*, 0.2% for *Necator americanus*, and 3.0% for *Strongyloides stercoralis* [27]. More recently, in a rural Alabama community—similar to the communities selected for this study—the prevalence was 34.5% for *Necator americanus* and 7.3% for *Strongyloides sterocoralis* [7], Although some studies have utilized soil samples for these helminths, none were conducted in the United States [13]. Nonetheless, as soil-transmitted helminths, they can also be ingested from the soil by children. Furthermore, the burdens of these parasites in soil have been associated with their prevalences in their sampling areas [14].

Ultimately, this study is the first to examine soil samples for an array of soil-transmitted helminths and unicellular parasites. The high contamination rates for *Blastocystis* spp. and *Toxocara* spp. as well as the detection of *Cryptosporidium* spp, *Giardia intestinalis, Ancylostoma duodenale, Trichuris trichiura, Necator americanus*, and *Strongyloides stercoralis* in soil may indicate their prevalence in their respective communities or serve as a source of infection.

### Relation of Environmental Contamination of Parasites to Poverty

The significant associations between the contamination rates of parasites and community poverty rates further confirm the well-established relationship between the risk of contracting gastrointestinal parasites and socioeconomic status [4]. Doni et al. (2015) found that the poor socioeconomic status of families, in addition to children’s behavior playing with soil, had the greatest association with the risk of parasitic infections for a cohort of children in Turkey [4].

However, the mediating factors in the relationship between parasites and poverty are not elucidated in our study. One such mediating factor is likely poor sanitation. Rural communities in the United States frequently lack access to municipal sanitation systems and instead rely primarily on septic tanks, which require maintenance and are vulnerable to overflow and back-up [4]. The increased risk of exposure to raw sewage can also increase the risk of infection with gastrointestinal parasites.

*Toxocara* spp. is a zoonotic pathogen that can be transmitted person-to-person and is not associated with poor sanitation. However, the results demonstrated a significant association and a strong correlation between environmental contamination and community poverty rate. Similar results were identified in a study of soil samples from public parks from the boroughs of New York City by Tyungu et al. (2020) [28]. In that study, the percentage of parks positive for *Toxocara* spp. was significantly associated with the median income of the borough. Furthermore, the burdens of *Toxocara* spp. eggs differed significantly with the highest burden in the borough with the lowest median income (Figure 5). These associations may be explained by the relationship between higher incomes and the ability to pay for veterinary check-ups and deworming.[28]

**Figure 5.**
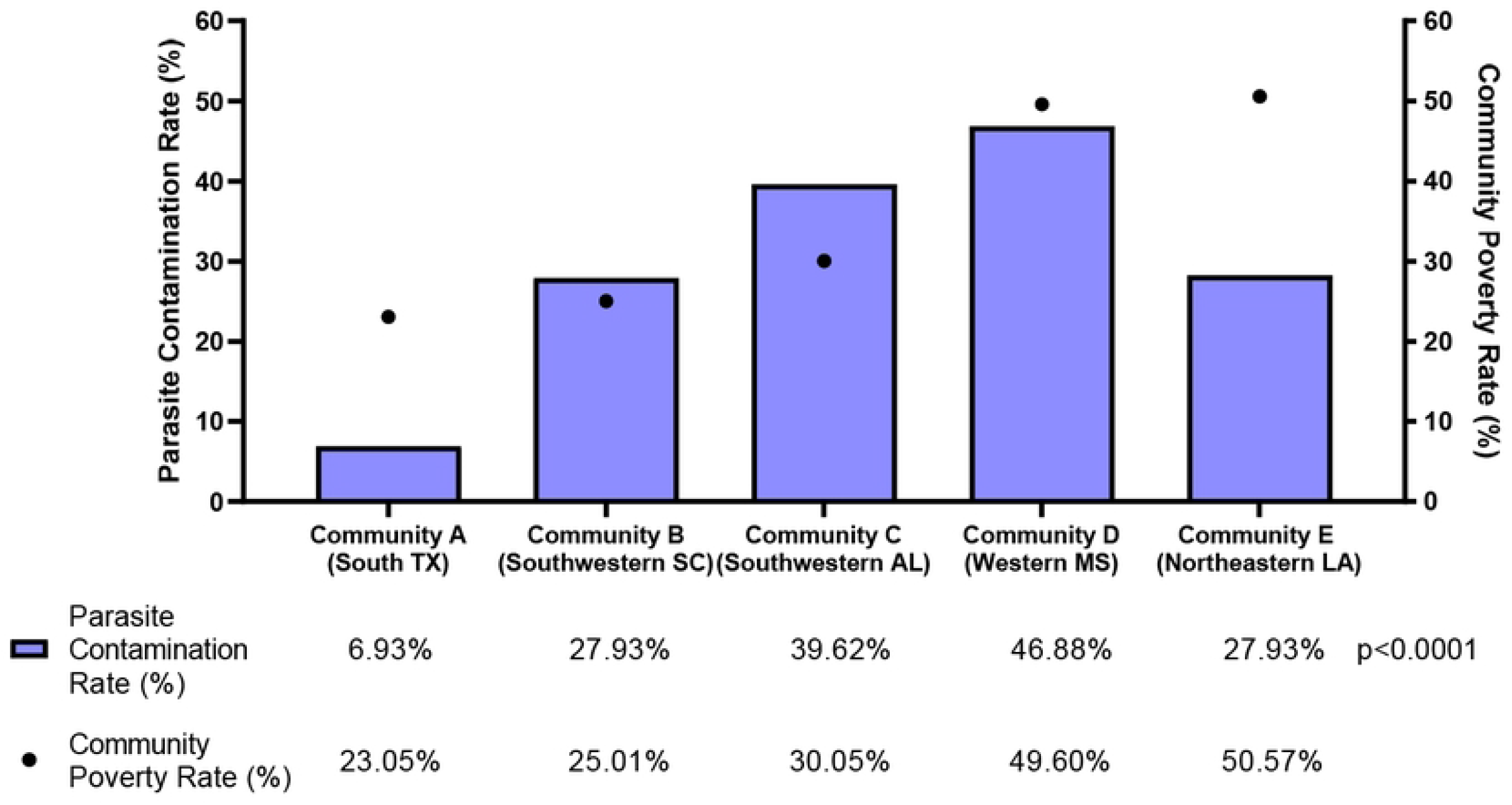
Normalized *Toxocara cati* burdens (fg/μL of DNA per kg of soil) by community. *Toxocara cati* burdens were significantly greater for communities with higher poverty rates (p=0.0011). Community poverty rates were obtained from the 2019 American Community Survey by U.S. Census Bureau.

Although our study demonstrated a relationship between parasites and poverty, it must be noted that poverty rate is not the only factor that influences the environmental contamination rates of parasites. A major limitation to environmental sampling is the heterogeneity of the occurrence of parasites in samples [29]. Furthermore, the contamination rates and burdens vary by soil type, as sandy soils allow greater parasite burdens than clay or silt [30].

### Limitations

While all attempts to maximize sample size per US state, there was limitations on collecting and processing dirt samples. Ideally, locations for sample collection should include areas of low poverty as a better representation of the link between poverty and environmental parasites. Although, this study focused on the at-risk populations in the United States. Also, while the primer and probes sets (Table 2) are specific for parasite DNA sequences, there is cross-reactivity noted for *Ancylostoma* primer/probe set that may detect the species *braziliense, caninum, ceylanicum*, and *tubaeforme* (https://blast.ncbi.nlm.nih.gov/Blast.cgi?PROGRAM=blastn&PAGE_TYPE=BlastSearch&LINK_LOC=blasthome). Several of the *Ancylostoma* species are zoonotic transmission, but do signify increase animal exposure, that maybe associated with rural and poverty conditions. Future studies will include a species-specific primer/probe set for *Ancylostoma duodenale*. The other parasites primer/probe sets are species specific at the time of this study completion.

The communities in this study are also located in different regions of the United States with different climates. Moreover, the samples were collected at different times of the year across the five communities. Temperature, rainfall, and relative humidity have been shown to affect the incidence of *Cryptosporidium* spp, among other parasites, and seasonal variations in parasite infections such as *Cryptosporidium* spp. and *Blastocystis* spp. have been identified [18]. Further work may attempt to elucidate the effects of some of these factors on parasite contamination rates and burdens, controlling for poverty rate. Alternatively, the relation between parasites and poverty rates may be further elucidated by controlling for climatic factors.

## Conclusion

Overall, several parasites were environmentally present in extreme low-income communities in the Southern United States. And despite high rates of poverty in all the communities studied, parasite contamination rates and burdens were associated with poverty rates. This indicates greater prevalence among human populations in communities with higher poverty rates and demonstrates greater potential for transmission in these communities.

## Data Availability

All data produced in the present work are contained in the manuscript.

## Contributors

CCB and RM conceptualized and designed the study. CCB, SY, DM, and RM designed the methods. CCB, SY, DM, LNH, RBI, MDB, and RM processed the samples. CCB, SY, and RM conducted the formal data analysis. The full data were verified by CCB, SY, and RM. SY and RM drafted figures with input from other authors. CCB, MEB, PJH, and RM provided oversight and leadership for the study. All authors approved the manuscript. All authors had full access to all the data in the study and had final responsibility for the decision to submit for publication.

## Acknowledgments

The authors would like to acknowledge and appreciate the organizers and people living in these Southern US communities for their support and willingness to cooperate with this study.

